# Cross-sectional analysis of Open payments for physicians at designated hemophilia centers in the US (2018-2020)

**DOI:** 10.1101/2023.03.07.23286934

**Authors:** Alyson Haslam, Vinay Prasad

**Affiliations:** Department of Epidemiology and Biostatistics, University of California San Francisco, 550 16th St, 2nd Fl, San Francisco, CA 94158

**Author notes:** **Corresponding author:** Dr. Alyson Haslam, Department of Epidemiology and Biostatistics, UCSF Mission Bay Campus | Mission Hall: Global Health & Clinical Sciences Building | 550 16th St, 2nd Fl, San Francisco, CA 94158. **Funding:** Arnold Ventures.

**Keywords:** hemophilia, conflict of interest, patient care

## Abstract

**Introduction:** Physicians who treat hemophilia, and especially directors at hemophilia centers, are in a position to be unduly influenced by payments from pharmaceutical companies who make costly hemophilia drugs. It is from this perspective that we analyzed payments made to physicians at hemophilia centers in the US, focusing on center directors.

**Materials and methods:** In a cross-sectional analysis we searched the CDC’s Hemophilia Treatment Center Directory for physicians (2022) and then abstracted general payments for physicians on Open Payments (2018-2020) and calculated one-year average payments. We searched academic websites to determine physician role (hemophilia center director, non-director, or non-center director).

**Results:** There were 420 physicians in the hemophilia physician directory – 270 physicians/professors, 103 directors of hemophilia centers, and 47 other directors. Directors of hemophilia centers had higher median one-year general payments, compared to other directors and physician/professors ($4910 vs $79 vs $87, respectively; p<0.0001). Takeda Pharmaceutical Company Limited, F-Hoffmann La Roche Ltd/Genentech, and Novo Nordisk have the largest hemophilia drug market share and were the three companies with the most payments to physicians.

**Conclusions:** High payments, especially among individuals who have responsibility over the success of hemophilia centers and clinics, may result in competition with the interest of the patients at these centers and clinics.

## Introduction

The Open Payment database was created under the Sunshine Act in 2013 in an effort to create more transparency in financial relationships between treating physicians and industry by mandating the reporting of all financial payments by drug, biological, device and medical supply companies to physicians. The database was created in part because of concern over conflict of interest influencing prescribing practices of physicians.

The issue of conflict of interest in the field of hemophilia is especially important since drugs for this condition are extremely expensive, with average treatment costs of $287,055, with most of the cost attributed to medication and clotting factor costs,(1) making this condition one of the most expensive to treat.(2) Industry payment amounts to oncologists/hematologists have increased since 2014, but fewer oncologists/hematologists received payments,(3) suggesting that conflict of interest may be greater among a select segment of oncology physicians.

Directors of medical centers are in a unique position to be able to influence patient care at a broad level, and because of their position, they may be targeted for industry funding. This may lead to conflict of interest between the interests of the patients and the interests of the medical center or clinic for which the director works.

We investigated physician-level payments among physicians who treat patients with hemophilia, and we examined whether there were differences in general payments between directors of hemophilia centers and non-directors.

## Methods

### Data and abstraction

We used the Centers for Disease Control and Prevention’s (CDC) Hemophilia Treatment Center Directory (https://dbdgateway.cdc.gov/HTCDirSearch.aspx) to compile a list of practicing hemophilia physicians in the US. The list includes name of physician, state, region, title, and name of treatment center. For each physician on the list, we used Open Payments (https://openpaymentsdata.cms.gov/) to find payments made to them by industry companies. We abstracted data for years 2018-2020. We abstracted data on general payment dollar amounts, number of general payments, general payment type, companies who made general payment, associated research funding, and number of payments for associated research funding. We calculated an average one-year general payment amount ($) and number of general payments for each physician by averaging years 2018-2020. We searched facility websites for title position for each physician. We classified physicians as physician/professor (including assistant and associate), director of hemophilia center, and other non-hemophilia leadership position (including director of fellowship, division director, etc.). For the companies with the top number of payments, we searched company websites for hemophilia drugs the companies made.

We used the CDC’s region classification, which is as follows: New England, Region I (Connecticut, Maine, Massachusetts, New Hampshire, Rhode Island, Vermont); New England, Region II (New Jersey, New York, and Puerto Rico), Mid-Atlantic (Delaware, Maryland (including Washington DC), Pennsylvania, Virginia, and West Virginia); Southeast, Region IV-North (Kentucky, North Carolina, South Carolina, and Tennessee); Southeast, Region IV-South (Alabama, Florida, Georgia, and Mississippi); Great Lakes (Indiana, Michigan, and Ohio); Northern States (Illinois, Minnesota, North Dakota, South Dakota, and Wisconsin); Great Plains (Arkansas, Iowa, Kansas, Louisiana, Missouri, Nebraska, Oklahoma, and Texas); Mountain states (Alaska, Arizona, Colorado, Idaho, Montana, New Mexico, Oregon, Utah, Washington, Wyoming); and Western States (California, Guam, Hawaii, and Nevada).

For the three companies with the highest number of payments to hemophilia physicians, we searched the Centers for Medicare and Medicaid Services Open Payments to find the amount of dollars paid out to hemophilia/oncology physicians. We used 2019 values because it was the most recent year before COVID changes were observed and because it aligns with the most recent estimate of market share for hemophilia drugs.

### Statistical analysis

We calculated median and mean dollar amounts and number of payments for physicians (total and by title (director vs not), region, and state. We looked at individual years (2018-2020) and an average of the three years. A Kruskal Wallace test was used to test for differences in medians, and a t-test was used to test for differences in means. We mapped median dollar amounts of general payments by state by using the ggplot2 and maps packages in R. All analyses were performed in R statistical software and Microsoft Excel. We used a 2-tailed alpha level of 0.05 for statistical significance. We used publicly available data and did not involve individual patient data, thus in accordance with 45 CFR §46.102(f), this study was not submitted for institutional review board approval. We adhered to STROBE reporting guidelines.

## Results

There were 420 physicians in the hemophilia physician directory – 270 physicians/professors, 103 directors of hemophilia centers, and 47 other directors. California had the highest number of hemophilia physicians (n=39), followed by Pennsylvania (n=33); Michigan (n=31); and Texas (n=28). Wyoming and Kansas had no hemophilia physicians. The median dollar amount for average one-year general payments was $156 (IQR: $10 to $6276), and the median one-year number of payments was two (IQR: 0.3 to 11). The highest average one-year general payment was $250045, and the highest number of general payments was 161. Eighty percent of hemophilia physicians received some amount of general payment (n=337), and 18% (n=77) received over $10,000 (Figure 1). The median one-year general payment was $0 for 2020 (IQR: $0 to $3143), $120 for 2019 (IQR: $0 to $6624); and $108 for 2018 (IQR: $0 to 4940). The median dollar amount of general payments per number of payments was $53 (IQR: $7.3 to $763). The median dollar amount of general payments per number of different companies paying a physician was $68 (IQR: $8.3 to $24393).

**Figure 1.**
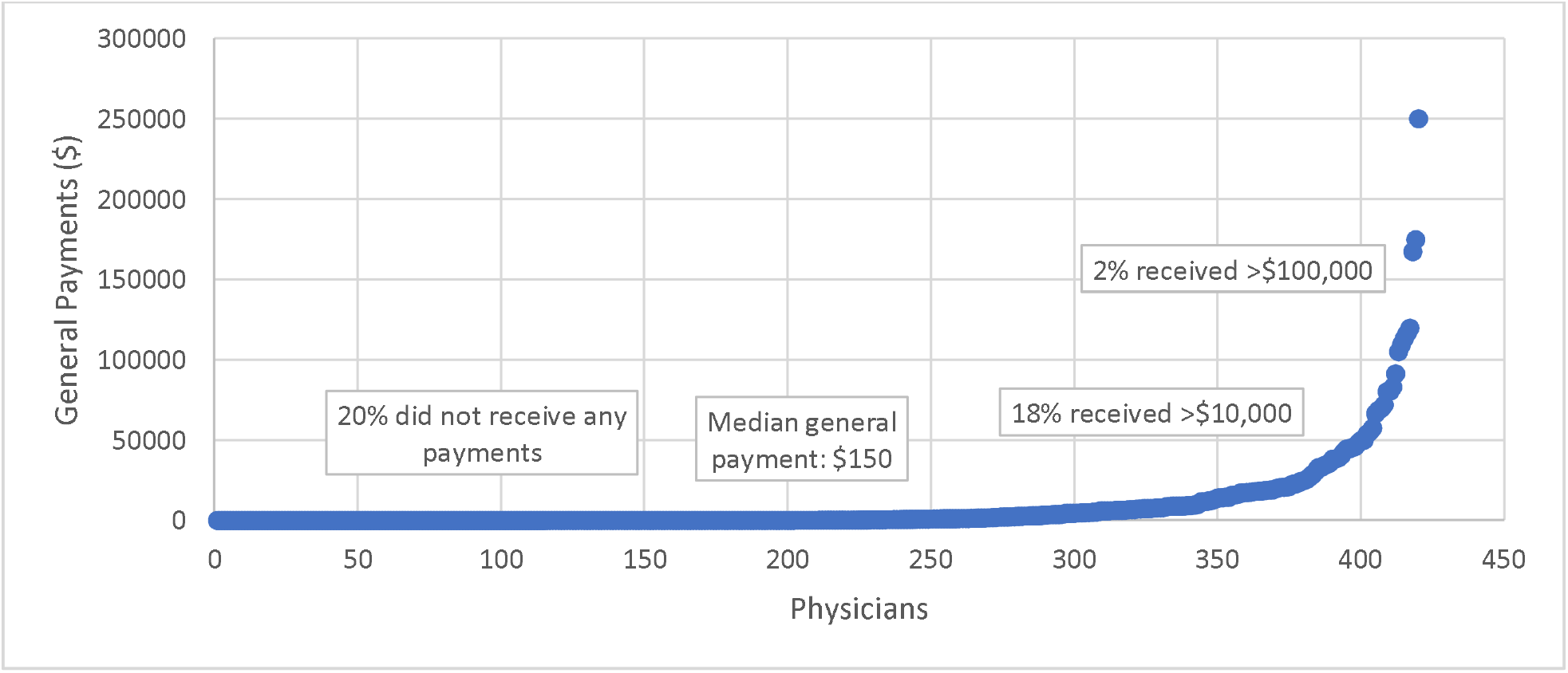
Average one-year general payment for hemophilia physicians in the US (2018-2020).

Directors of hemophilia centers had higher median one-year general payments, compared to other directors and physician/professors ($4910 vs $79 vs $87, respectively; p<0.0001; Table 1). Similarly, we found this to be true for 2018 ($980 vs $67 vs $76; p<0.0001), 2019 ($5907 vs $37 vs $64; p<0.0001), and 2020 ($2449 vs $0 vs $0; p<0.0001). Directors of hemophilia centers also had a higher median number of general payments (6 vs 1 vs 1; p<0.0001), and directors had a higher dollar amount per payment compared to physicians or other directors ($475 vs $36 vs $40; p<0.0001). The percentage of physicians who received over $10,000 in general payments was higher for directors of hemophilia centers, compared to non-directors (34% vs 13%; p=0.0001).

**Table 1.** Median general payments for hemophilia physicians, by leadership status (2018-2020)

|  | Physician/professor<br>(n=270) | Director of<br>hemophilia<br>center<br>(n=103) | Other leadership<br>(n=47) | p-value<sup>1</sup> |
| --- | --- | --- | --- | --- |
| **Average one-year**<br>**general**<br>**payments, 2018-**<br>**2020** |  |  |  |  |
| Mean (SD) | \$5819 (16275) | \$19027 (38707) | \$6526 (18417) | <0.0001 |
| Median (IQR) | \$87 (6-2948) | \$4910 (71-18198) | \$79 (6-2635) | <0.0001 |
| **Average number**<br>**of general**<br>**payments, 2018-**<br>**2020** |  |  |  |  |
| Mean (SD) | 8 (16) | 18 (29) | 5 (12) | <0.0001 |
| Median (IQR) | 1 (0.3-3) | 6 (1-19) | 1 (0.3-3) | <0.0001 |
| **General**<br>**payments, 2018** |  |  |  |  |
| Mean (SD) | \$6225 (19750) | \$21519 (50016) | \$7944 (21614) | <0.0001 |
| Median (IQR) | \$76 (0-1928) | \$980 (25-19543) | \$67 (0-3428) | <0.0001 |
| **Number of**<br>**general**<br>**payments, 2018** |  |  |  |  |
| Mean (SD) | 9 (22) | 21 (35) | 7 (14) | <0.0001 |
| Median (IQR) | 1 (0-9) | 7 (1-24) | 1 (0-4) | <0.0001 |
| **General**<br>**payments, 2019** |  |  |  |  |
| Mean (SD) | \$8413 (27792) | \$23259 (46130) | \$7179 (23341) | <0.0001 |
| Median (IQR) | \$64 (0-1821) | \$5907 (19-21438) | \$37 (0-4551) | <0.0001 |
| **Number of**<br>**general**<br>**payments, 2019** |  |  |  |  |
| Mean (SD) | 10 (23) | 25 (40) | 7 (17) | <0.0001 |
| Median (IQR) | 1 (0-8) | 8 (1-24) | 1 (0-6) | <0.0001 |
| **General**<br>**payments, 2020** |  |  |  |  |
| Mean (SD) | \$3200 (9194) | \$12304 (23087) | \$4453 (12955) | <0.0001 |
| Median (IQR) | \$0 (0-470) | \$2449 (0-11455) | \$0 (0-408) | <0.0001 |
| **Number of**<br>**general**<br>**payments, 2020** |  |  |  |  |
| Mean (SD) | 3 (7) | 9 (16) | 3 (7) | <0.0001 |
| Median (IQR) | 0 (0-2) | 2 (0-11) | 0 (0-2) | <0.0001 |

1. Kruskal Wallace for medians and t-test for means.

We found that there were significant differences in median one-year general payments, by region (p=0.0003). The regions with the highest median general payment were the Southeast ($3295; Georgia, Alabama, Mississippi, Florida; region 5), followed by California/Nevada ($1206; region 10). All other regions were under $200. We also found differences in general payments, by state (p=0.003). Among all physicians included in the CDC’s hemophilia directory, Kentucky was the state with the highest median general payment ($22513), followed by Louisiana ($22250), Mississippi ($11670), and Iowa ($8542). When only including physicians in the top 50% of general payments, Connecticut ($49959), Oregon ($35745), and Kentucky ($22513), and Louisiana ($22250) were the states with the highest median general payments (Figure 2).

**Figure 2.**
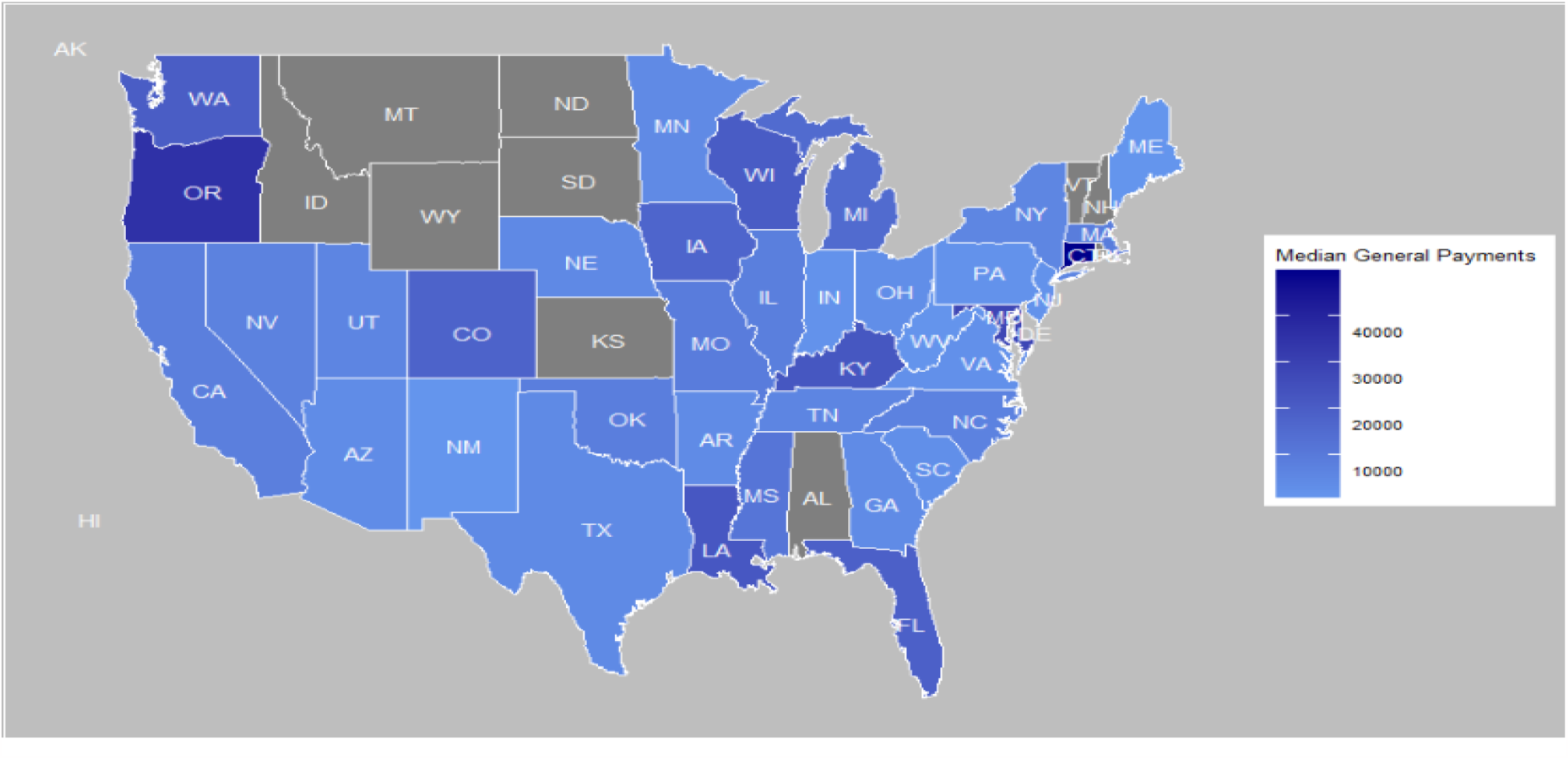
Median 1-year average general payments (2018-2020) for hemophilia physicians in the top 50^th^ percentile of general payments, by state.

The companies with payments to the greatest number of physicians were Genentech/F-Hoffmann La Roche (n=70), Novo Nordisk (n=63), and Takeda/Shire (n=60) for 2020; Genentech/F-Hoffmann La Roche (n=127), Shire (n=84), and Novo Nordisk (n=77) for 2019; and Genentech/F-Hoffmann La Roche (n=142), Shire (n=89), and Novo Nordisk (n=79) for 2018. With the exception of Novo Nordisk during 2018, directors were more likely than physicians/professors to receive money from these mentioned companies. Genentech paid hemophilia/oncology physicians $4526794.12 in 2019, Takeda paid $995117.80, and Novo Nordisk paid $705799.56 to hemophilia/oncology physicians.

The hemophilia drugs that these companies sell are: Novo Nordisk makes antihemophilic factor (recombinant), glycopegylated-exei, antihemophilic Factor (Recombinant), coagulation Factor VIIa (recombinant), coagulation Factor IX (recombinant), GlycoPEGylated, and coagulation Factor XIII A-Subunit (recombinant); Roche/ Genentech makes emicizumab-kxwh; Takeda (acquired Shire in 2019) makes the orphan drug antihemophilic factor (recombinant), PEGylated and factor eight inhibitor bypassing activity.

## Discussion

Among physicians listed on the CDC’s hemophilia physician directory, we found that 13% of non-directors at hemophilia centers received over $10,000 in general payments and 34% of directors of hemophilia centers received over $10,000, with the median amount received being $4910 per annum for directors. Furthermore, directors received a greater number of payments, and the dollar amount for each payment was higher than non-directors.

One concern is that general payments from industry could lead to higher health care costs with no concomitant benefit for the patients.(4) Previous analysis show that the receipt of higher general payments is associated with a higher likelihood of prescribing costlier drugs when there are less expensive and perhaps equally effective formulations available.(5) Oncologists who received payments from a drug company were more likely than physicians not receiving payment from that company to prescribe a drug made by that company.(6) While it was beyond the scope of our project to assess prescribing practices of directors of hemophilia centers, there is cause for concern that the higher industry payments to directors could bias treatment decisions made at hemophilia treatment centers, thus putting the interests of the centers above those of the patients who are patients at the centers.

Median general payments vary by medical specialty, with medical specialists receiving more than primary care, surgical, hospital-based, and obstetrics/gynecologist physicians.(7) The amount of general payments for hemophilia physicians was higher than most medical specialties, but lower than medical oncologists, who received a median of $791 in general payments (2019).(3) While the amount of general payments among hemophilia physicians in our study was lower than the previously reported amount among medical oncologists, it is not directly comparable because our median estimate also included physicians who did not receive any general payments and because the CDC’s list of hemophilia physicians includes non-oncology/hematology physicians (n=15). When only including physicians who received any payment, the median general payment was $916, which is higher than the previously reported amount medical oncologists receive. The higher median payments of physicians who treat hemophilia parallels the exorbitant drug costs for this condition.

The global hemophilia drugs market size was valued at $11.8 billion in 2019 and is estimated to reach $15.8 billion by 2026.(8) Takeda Pharmaceutical Company Limited, F-Hoffmann La Roche Ltd (parent company of Genentech), and Novo Nordisk have the largest market share of hemophilia drugs, and they were also the three companies with payments to the highest number of physicians. The trend toward prescribing newer and costlier drugs(9) will likely drive these figures even higher in future years. Yet, payments from these three companies to hematologists/oncologists accounted for about 0.05% of the hemophilia market size for 2019.

There are at least 3 strengths and 4 weaknesses to our analysis. First, we searched a comprehensive set of hemophilia experts, as endorsed by the CDC. Second, we geographically mapped physician payments to show both state and regional trends in physician payments to hemophilia physicians. Third, we averaged several years of data to limit the year-to-year variability.

There are several limitations to our analysis. First, our analysis was restricted to physicians that were included on the CDC’s hemophilia center physicians, and there are other physicians who treat patients with hemophilia who were not on the list. Hence, our results do not apply to all physicians who treat hemophilia. Second, our classification of leadership status was based on the online information we found from the hospital/institution’s website where the physician was employed. These results may not have been up to date or their leadership status may not have been displayed on the website. This would have resulted in an incorrect categorization in leadership position. However, it is likely that this would have resulted in non-differential misclassification. Third, our results may not be broadly representative because of changes in payments during COVID (2020). To get a more general representation of payments, we took an average of the most recent three years of data. Fourth, when looking at median payments by state, variability in the number of practicing hemophilia physicians meant that individual data points had stronger influence on median payments. Median payments by region may be a better overall indicator of general payments since the number of physicians were higher for each and had less variability when compared to states.

## Conclusion

In conclusion, physicians who treat patients with hemophilia receive a higher amount of industry payments compared to other medical specialties. Further, directors of hemophilia centers, who are in positions to influence clinical care at a broad level, including crafting local treatment algorithms, receive much higher amounts of industry renumeration compared to non-directors. High payments, especially among individuals who have responsibility over the success of hemophilia centers and clinics may result in competition with the interest of the patients at these centers and clinics.

## Data Availability

All data produced are available online at CMS Open Payments (https://openpaymentsdata.cms.gov/) and Centers for Disease Control and Prevention (CDC) Hemophilia Treatment Center Directory (https://dbdgateway.cdc.gov/HTCDirSearch.aspx)

## Abbreviations

CDC: Centers for Disease Control and Prevention

## Data sharing statement

All data are publicly available on US government websites.

## Author Contribution

VP conceptualized study design and edited manuscript; AH abstracted data, conducted data analysis, and drafted manuscript.

